# Mexico’s surge of violence and COVID-19 drive life expectancy losses 2015–2021

**DOI:** 10.1101/2024.05.07.24306982

**Authors:** Jesús-Daniel Zazueta-Borboa, Paola Vázquez-Castillo, Maria Gargiulo, José Manuel Aburto

**Author notes:** Corresponding author: JDZB (;).

## Abstract

**Background:** Life expectancy at birth in Mexico has stagnated since the early 2000s. As the COVID-19 pandemic hit, Mexico experienced sizable excess mortality, albeit with large regional variation. We aimed to assess the contribution of violence, COVID-19, and causes of death amenable to healthcare to life expectancy changes between 2015 and 2021 in Mexico.

**Methods:** We used administrative mortality data by causes of death, and adjusted population exposures from the National Population Council. We applied demographic decomposition methods to assess life expectancy changes at the subnational level, by year and sex.

**Findings:** Life expectancy between 2015 and 2019 declined from 71.8 to 71.1 years for males and stagnated at 77.6 years for females. Violence among young males accounted for 54.3% of life expectancy losses. Between 2019 and 2020, life expectancy decreased by 7.1 and 4.4 years for males and females, respectively. COVID-19 deaths accounted for 55.4% (males) and 57.7% (females). In 2021, male life expectancy stagnated at 64.1 years due to reductions in deaths due to amenable diseases but continued increasing for females by 0.44 years mainly due to reductions in COVID-19 deaths.

**Interpretation:** We document large variations in life expectancy losses across Mexican states, which are associated with preexistent high levels of violence, and socioeconomic disadvantages across geographical areas. Our results serve as a reminder that violence has negative health implications for both sexes and that COVID-19 affected more socially disadvantaged states.

**Funding:** Support from the Netherlands Interdisciplinary Demographic Institute-KNAW, AXA Research Fund, Economic and Social Research Council, and European Union’s Horizon 2020 Research and Innovation.

**Research in context.:** *Evidence before this study:* We searched for studies in English and Spanish that analyzed life expectancy losses in Mexico before and during the COVID-19 pandemic in PubMed. Most studies that assessed life expectancy during the COVID-19 rely on all-cause mortality and indirect demographic methods. We also identified studies on causes of death, those focused on age-standardized mortality or excess deaths, but as of April 2024, we did not find articles assessing the impact of multiple causes of death on life expectancy.

*Added value of this study:* To our knowledge, this is the first study to assess the impact of different causes of death on life expectancy before and during the COVID-19 pandemic at the subnational level and by sex in Mexico. We focus on the main causes of death including COVID-19, homicides, and causes amenable to health care (e.g. diabetes). Our findings reveal that before the COVID-19 pandemic (2015–2019) life expectancy decreased for males and remained the same for females. During 2019–2020 life expectancy decreased sharply for both males and females, while in the subsequent years (2020–2021), life expectancy roughly remained the same for males, and continued decreasing for females. Most of the life expectancy losses before the pandemic for males were due to violence and homicides, while since 2020 they were due to COVID-19, but diabetes and causes of death amenable to health care also contributed to reduced life expectancy. Life expectancy losses were unevenly distributed at the subnational level, states from southern and central Mexico experienced the largest life expectancy losses compared to states from north of Mexico.

*Implications of all the available evidence:* This study contributes to understanding life expectancy changes before and during the COVID-19 pandemic years. By quantifying life expectancy losses we uncover the unequal and devastating impact of the pandemic at the subnational level in Mexico. Moreover, our results highlight the continued failure on reducing homicides and violence in the country.

## Introduction

Latin America is one of the most violent regions in the world. Several countries, including Mexico, have experienced unprecedented waves of violence over the course of the 21^st^ century, (1) with sizable impacts on population health, life expectancy, and lifetime uncertainty.(2) In 2017, Latin America accounted for 7.8% of the global population, but 37% of global victims of intentional homicides, with a homicide rate of 17.2 per 100,000 people; the highest recorded in the region since 1990.(3)

Alongside the unprecedented rise in homicides, Latin America has been heavily affected by the COVID-19 pandemic.(4) The lack of timely policy responses, unequal health care access, and high prevalence of cardiovascular disease and comorbidities led to a high burden of excess mortality in the region.(5) For some Latin American countries, life expectancy losses at the beginning of the pandemic ranged from 3 (Chilean females) to 10 years (Peruvian males).(4, 6, 7) In Mexico, life expectancy losses in 2020 were 2.7 and 3.5 years for females and males respectively, with large regional variation.(7) It is unknown, however, how increased violence, causes of death amenable to healthcare (e.g., respiratory diseases, and some neoplasms), and the COVID-19 pandemic have affected life expectancy in Mexico in more recent years.

In Mexico, through 2015, violence contributed to life expectancy declines among males, and stagnation among females.(8, 9) The homicide rate among males increased rapidly from 22 to 43 deaths per 100,000 people between 2006 and 2012. Between 2012 and 2014, it decreased slightly, but in 2015, it started to increase again, reaching its maximum in 2019 with a homicide rate of 75 deaths per 100,000 people. (See Supplemental Figure 9). In the period 2006–2019, there were 325,884 homicides recorded, of which 88.8% corresponded to males.(10) The increase in homicide rates is the result of public policies that allowed military interventions to mitigate drug cartels’ operations that started in December 2006, the beginning of the so-called “war on drugs”.(8, 9)

Another important public health challenge for Mexico is the increase in diabetes-attributable mortality, which has also contributed to life expectancy stagnation.(9) Between 2000–2018, the prevalence of diabetes increased from 7.5% to 10.3%, (11) and the number of deaths attributed to diabetes increased by 77% between 1990 and 2017. In total, diabetes accounted for almost a million deaths (948,532) between 2010 and 2019.(10) After diabetes-attributable mortality and violence, some causes of death amenable to healthcare, i.e., those causes that should not result in death when effective and pertinent medical care is provided, have also increased. Among them are infectious and respiratory diseases, and some types of cancer, such as sex-specific cancers.(12) Other causes amenable to healthcare, such as infectious and respiratory diseases, contributed to increases in life expectancy between 2000– 2015.(8, 9)

This article makes three main contributions. First, we document the contribution of violence, diabetes, causes of death amenable to healthcare, and the COVID-19 pandemic to life expectancy changes in the period 2015–2021. This is the first study to look at the contribution of different causes of death to life expectancy in Mexico in this period. Second, we contribute to the existing literature on violent mortality, by looking at the most recent years, which have been characterized by increasing homicides among males and females between 2015–2019. Third, this analysis contributes to our knowledge of the impact of different causes of death on life expectancy changes before and during the COVID-19 pandemic across Mexican states.

We analyzed the contribution of different age groups and causes of death to life expectancy changes for males and females between 2015 and 2021, we focused on changes before (2015–2019) and during the pandemic years (2019–2020, 2020–2021). This framework allows us to thoroughly assess the contribution of amenable diseases, diabetes, and violence to life expectancy losses before and during the pandemic years.

## Data & Methods

### Data

We retrieved data from different sources. Death certificates (2015, 2019, 2020, and 2021) were downloaded from vital statistics files from the National Institute of Statistics.(10) These files contain information on causes of death, sex, and the age at the time of death. We used this data source to obtain the proportion of causes of death in each age group, by sex and year. From the National Population Council, we retrieved the population at risk and the number of deaths. We used the mid-year population estimates and deaths.(13) These estimates are corrected for completeness, age misstatement, and international migration. With both datasets, we constructed age-specific death rates by sex and state for the years 2015, 2019, 2020, and 2021.(13)

We focus our analysis on the main causes of death that previously contributed the most to life expectancy stagnation between 2005 and 2015 and COVID-19. This is to assess their impact after 2015 and their dynamics during the COVID-19 pandemic. We grouped deaths into six main categories: i) homicides and violent deaths, ii) COVID-19, iii) diabetes, iv) causes amenable to healthcare, v) external causes of death, and vi) all other causes. Analyzing COVID-19 and causes amenable to health care separately allows us to distinguish those causes that were primarily driven by SARS-CoV-2 from those that would have occurred in a non-pandemic year. We classified deaths using the International Classification of Diseases 10^th^ revision (ICD-10). The ICD-10 code groupings are presented in Supplemental Table 1.

## Methods

We first computed age-specific death rates by five-year age groups for each state and for the national level by sex. Afterward, we constructed period life tables at the national level and for all 32 Mexican states by sex for the period 2015–2021 using standard demographic techniques.(14) The main mortality outcome from this analysis was life expectancy at birth (e0). We later used the continuous-change decomposition method(15) to assess changes in e0 for the periods: 2015– 2019, 2019–2020, 2020–2021 using the R package “DemoDecomp”.(16) In addition, to contextualize mortality trends, we computed age-standardized death rates at national and subnational levels for all-cause and cause-specific mortality to draw trends from 2000-2021, using the 2010 Mexican population as standard (See Supplemental Figures 8-12) and trends in e0 at the national level and states (Supplemental Figure 7). Our analysis at the subnational level is stratified by regions, in Supplemental Figure 15 we show the map with states and regions of Mexico.

## Results

Figure 1 displays the age-specific and cause-of-death contribution to life expectancy changes by sex before the pandemic (2015–2019) and in two pandemic years (2019–2020 and 2020–2021). Between 2015 and 2019, life expectancy decreased by 0.7 years, from 71.8 years to 71.1 years for males, and stagnated with a slight increase of 0.1 years, from 77.5 to 77.6 years for females. The decline in life expectancy among males occurred mostly in ages 20–39 due to homicides and violence (54.3%). During the first year of the pandemic (2019–2020), life expectancy decreased by 7.1 years, from 71.2 years to 64.1 among males, and decreased by 4.5 years from 77.6 years to 73.1 years among females. Most of the decrease in life expectancy was attributed to COVID-19 deaths in adults ages 60 and over, accounting for 34.4% and 41.3% of life expectancy losses for males and females, respectively. In the second year of the pandemic (2020–2021), male life expectancy stagnated with a slight increase of 0.1 years from 64.1 years to 64.2 years, mostly due to improvements in diabetes and causes amenable to healthcare in individuals aged 60 and over. Conversely, for females, life expectancy continued to decrease by 0.4 years, from 73.1 years to 72.7 years. For females, COVID-19 contributed to decreased life expectancy at all ages, offsetting progress in diabetes after age 50.

**Figure 1.**
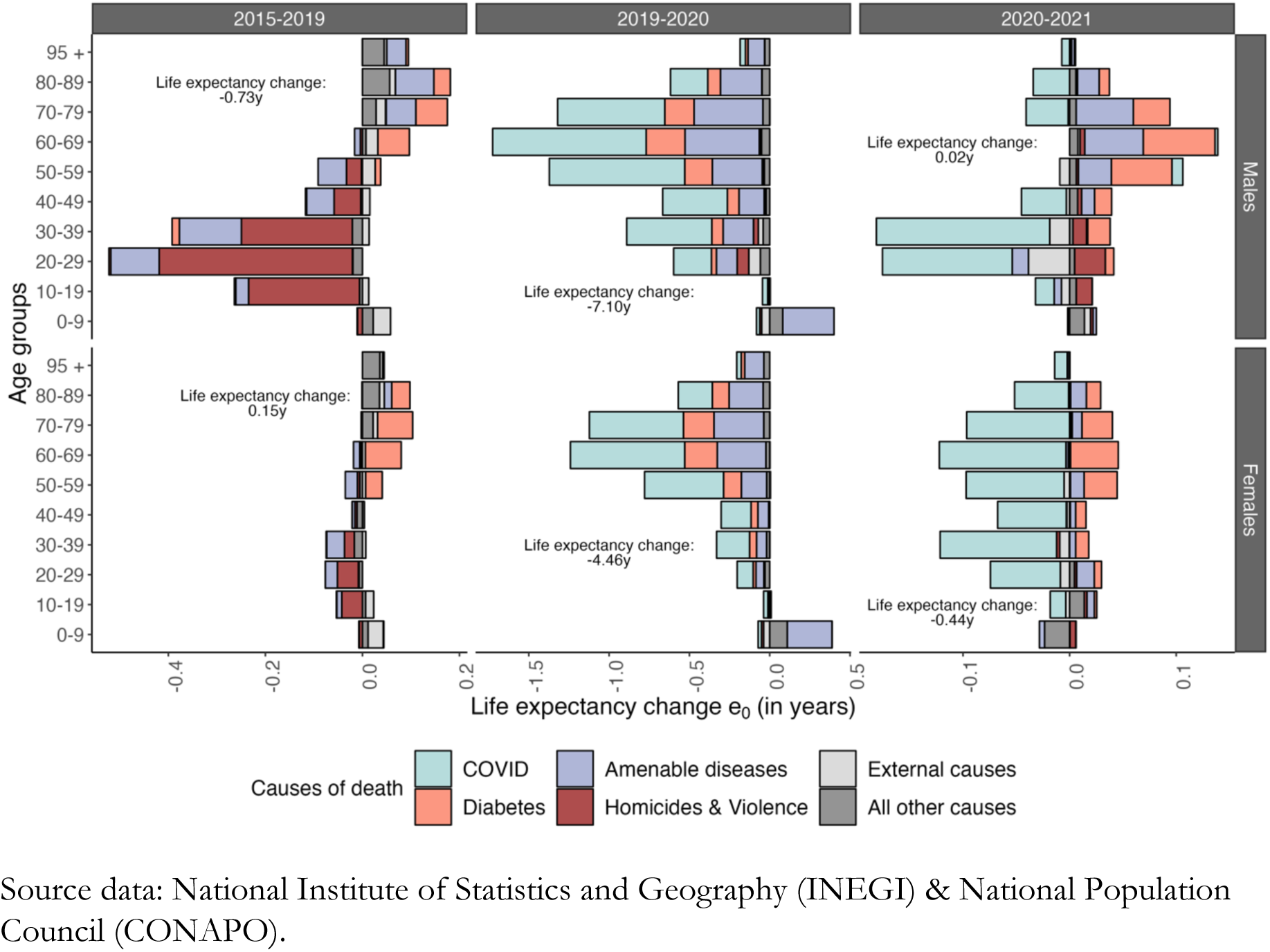
Cause-specific contributions to life-expectancy changes, by age, sex, and period, 2015–2021.

We observed large variations in life expectancy losses across Mexican states before and during the pandemic. Supplemental Figure 1 shows levels and changes in life expectancy at birth in 2019 and 2021 by sex and across Mexican states and the number of years of life expectancy losses (Supplemental Figure 2). Figures 2–5 show the total contribution of violence and homicides (Figure 2), COVID-19 (Figure 3), diabetes (Figure 4), and causes amenable to healthcare (Figure 5) to life expectancy changes at the state level by sex over the period 2015–2021. In the Supplementary Material, we present the age-specific contribution to life expectancy at the state level at the different periods (See Supplemental Figures 3-6).

**Figure 2.**
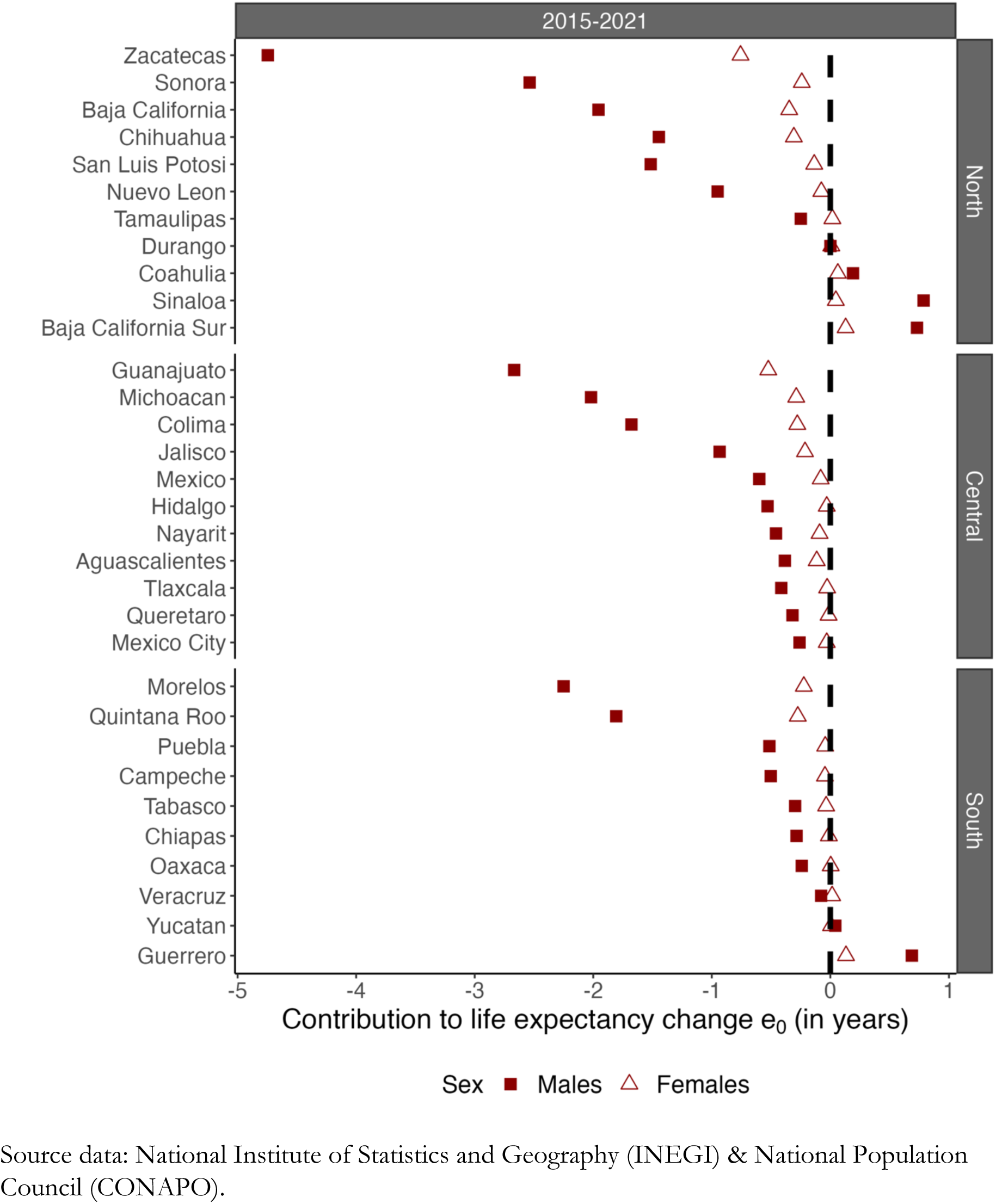
The contribution of homicides and violence to life expectancy changes between 2015–2021, by state and sex.

Between 2015 and 2021, homicides and violence contributed to reductions in life expectancy among males in 26 out of 32 states (Figure 2). The northern and central regions of Mexico are the most affected, with higher contributions of homicides and violence to changes in life expectancy. For females, homicides and violence contributed to a modest reduction in life expectancy at birth in 12 states. We observed concordance between the states with a higher contribution of homicides and violence for males and females, with a lower contribution for females. In Supplemental Figure 3, we show that among males in states of the northern region of Mexico for the period 2019–2020, homicides and violence contributed to reduced life expectancy at birth.

Across all Mexican states, COVID-19 contributed to life expectancy losses between 2019 and 2021, with regional variation (See Figure 3). For both males and females, the largest life expectancy losses occurred in central and southern Mexico. On average, the impact of COVID-19 on life expectancy changes was much smaller in northern Mexico. Supplemental Figure 4 shows that COVID-19 contributed positively to life expectancy changes (2020–2021) across some states in northern Mexico, while it continued contributing to reductions in life expectancy in the central and southern regions.

**Figure 3.**
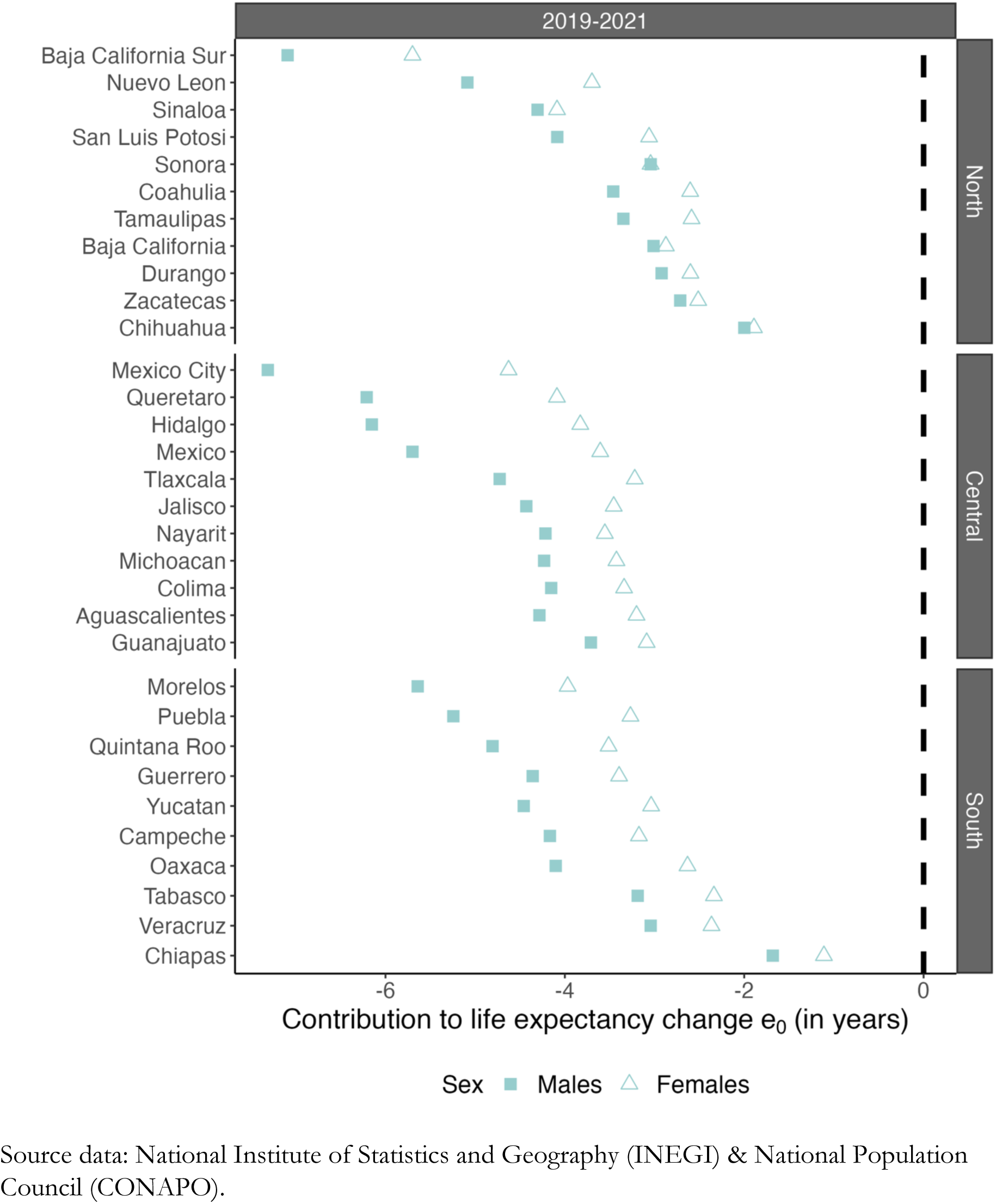
The contribution of COVID-19 to life expectancy changes between 2019–2021, by state and sex.

Mortality due to diabetes had a strong regional variation (See Figure 4). For half of the states in the northern region, diabetes contributed to increasing life expectancy at birth for both males and females, while it contributed to reductions in the southern and central regions. When looking at the contribution of diabetes to changes in life expectancy at birth over time (See Supplemental Figure 5), we observed that between 2015 and 2019, the changes in diabetes mortality contributed to increasing life expectancy at birth in the north and central regions, but not in the south of Mexico. However, during the pandemic 2019–2020, diabetes contributed to reductions in life expectancy at birth across all states, and between 2020 and 2021 reductions in diabetes-related deaths contributed to increasing life expectancy at birth in all states.

**Figure 4.**
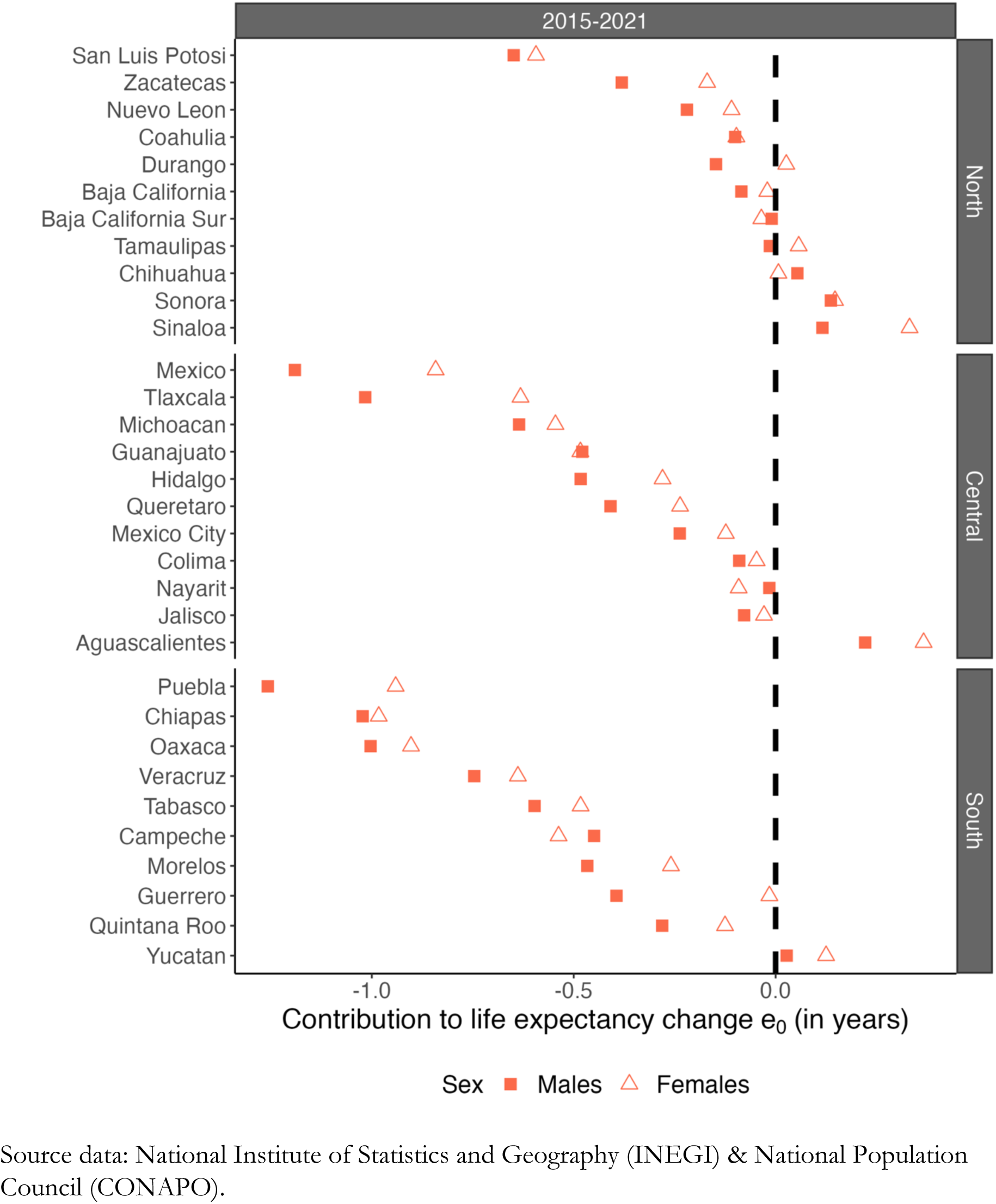
The contribution of diabetes to life expectancy changes between 2015–2021 by state and sex.

Causes of death amenable to healthcare contributed to reduced life expectancy between 2015– 2021 (See Figure 5). However, these causes contributed negatively to life expectancy changes for all regions. In south Mexico, these causes had a larger contribution to life expectancy losses, compared to north and central. This pattern was observed for both males and females. As with the contributions of COVID-19, we observed that in northern Mexico, causes amenable to healthcare contributed to increasing life expectancy (2020–2021) for both males and females, while central improvements are more modest, and in the south continued contributing to reducing life expectancy (See Supplemental Figure 6).

**Figure 5.**
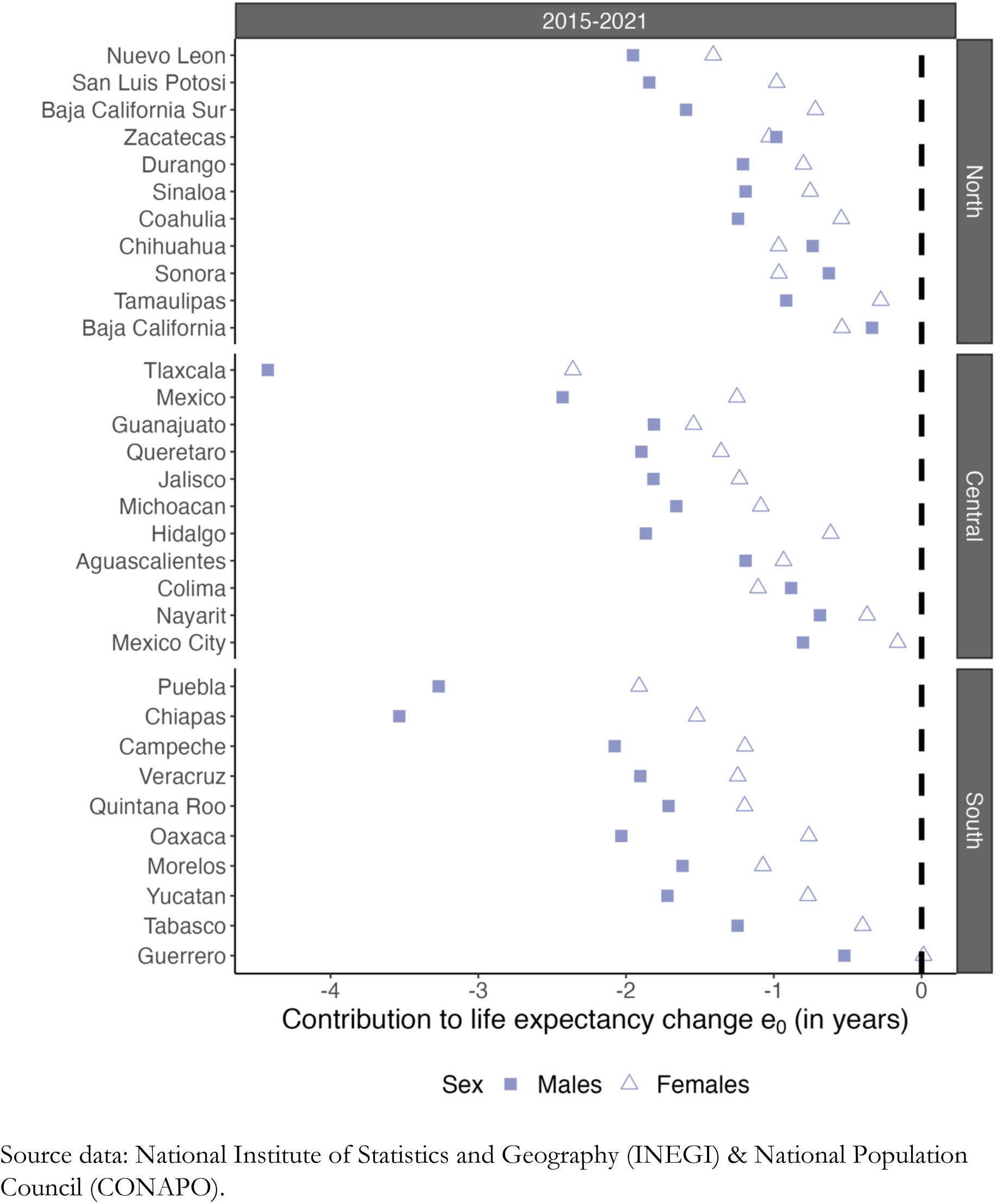
The contribution of causes amenable to healthcare to life expectancy changes between 2015–2021 by state and sex.

## Discussion

### Summary of results

Before the COVID-19 pandemic (2015–2019), male life expectancy at birth decreased by 0.8 years, from 71.8 years to 71.1 years, and female life expectancy at birth stagnated, at around 77.5 years. Violence among males aged 20–39 years accounted for 54.3% of life expectancy losses in the period. Between 2019–2020, life expectancy decreased by 7.1 years and 4.4 years at the national level for males and females, respectively. In 2021, male life expectancy at birth stagnated at 64.1 with improvements in diabetes at older ages. However, life expectancy continued decreasing for females resulting in losses of 0.44 years due to COVID-19 at all ages. We capture large variability in life expectancy losses across Mexican states ranging from 2.6 years for males in Tamaulipas (northern region) to 10.1 years for males in Tlaxcala (central region).

### Interpretation of results

Levels of violence in Mexico have increased since 2006. The violence that was once concentrated in the north of Mexico and primarily affected young males (8, 9, 17) has intensified and spread across the country. Our analysis reveals that, after 2015, the contribution of homicides and violence to life expectancy losses spread across all Mexican states. As has been observed in previous years, violence had a higher negative impact on male life expectancy than female life expectancy.(8, 9) However, between 2015–2021, violence contributed modestly to decreases in female life expectancy. A contribution of this nature had only previously been observed in states with extremely high levels of violence, such as Chihuahua in 2005–2009.(9) Furthermore, we observe that nationally and in almost all Mexican states, the number of deaths due to homicides and violence among females has increased over time (2000–2021) (see Supplemental Figure 9), but specifically in states from the central region such as Colima, Guanajuato, and Mexico City.

While previous studies on the relationship between violence and life expectancy have focused on males because they are killed violently more often than females, our results serve as a reminder that violence has negative health implications for both sexes. The changing nature of the contribution of violence to changes in female life expectancy is consistent with other research that has found that the nature of lethal violence against women has changed since 2006. (18) For example, while lethal violence against women was once characterized as deaths that occurred inside the home by someone the victim knew, violence against women in Mexico now has an increasingly public nature, with more women being killed in public places and by strangers.(19)

Many of these deaths form part of Mexico’s ongoing feminicide crisis, whereby women and girls are killed intentionally because of their gender. Public health measures aimed to curb the spread of COVID-19, such as lockdowns, may have also resulted in increased violence against women and girls, although this can also be difficult to quantify due to reporting challenges (20).

During 2020 and 2021, Mexico was severely impacted by the COVID-19 pandemic with variable effects across Mexican states. Life expectancy losses exceeded more than 2 years for both sexes across states. To put this number in perspective, in Italy and Spain, two countries that were epicenters of the COVID-19 pandemic, life expectancy losses in 2020 ranged from 1.0 years among Italian females to 1.5 years among Spanish females.(21) In the United States, the country with the second highest number of COVID-19-related deaths, life expectancy losses were 1.7 years and 2.2 years for females and males respectively,(21) although with large differences across ethnic groups.(22, 23) Moreover, in most European countries, life expectancy at birth in 2021 started recuperating, but levels are still lower than those reported in 2019.(24) Among Latin American countries, few studies have examined life expectancy losses at the subnational level. For Chile, life expectancy decreased by 1.8 years for males and 1.3 among females in 2020, but poorer urban municipalities were the most affected.(25) In Brazil, life expectancy decreased in 2020 and the Amazonian and northern regions experienced the largest losses. Overburdened healthcare facilities, including insufficient hospital beds and disruption in primary care services are potential explanations for regional differences and continued life-expectancy losses in Brazil in 2021.(6)

For Mexico, the largest losses in life expectancy at birth between 2019 and 2021 may be explained by preexistent comorbidities across socioeconomic groups. Sociodemographic and health profiles of individuals who died due to COVID-19 provide corroborating evidence for this hypothesis. Preexistent health risk factors among the population, such as diabetes and obesity, seem to be one of the reasons why Mexico was so heavily affected by COVID-19.(26, 27) However, studies using individual-level data on patients who tested positive for COVID-19 highlight that, after controlling for health risk factors, individuals’ socioeconomic characteristics were important in determining mortality outcomes.(28–30) For example, the probability of dying due to COVID-19 among individuals in the lowest income quantile was five times the probability of dying as those in the highest decile, even after accounting for health risk factors.(29) Individuals’ socioeconomic conditions were important for coping with lockdowns and mitigating virus spread. For example, having limited financial resources and depending on emergency government assistance made it harder for individuals to adhere to isolation measures.(30) This may also help explain why the case fatality rate among the indigenous population was 64% higher than among non-indigenous people.(26)

We observed large variability in life expectancy losses between 2019 and 2021 across Mexican states. Moreover, for all states and both sexes, life expectancy at birth in 2021 was much lower compared to 2019 levels (Supplemental Figure 1). In general, we observed that between 2019 and 2020, the impact of COVID-19 on life expectancy was similar across states. In 2020 and 2021, we observed recoveries from COVID-19 in states in northern Mexico, while states in the central and southern regions continued to experience life expectancy losses due to COVID-19 (Supplemental Figure 3). These regional differences seem to be driven by sociodemographic characteristics and health inequalities across states.(28, 30) Northern Mexico is characterized by lower levels of poverty and lower levels of social vulnerability index (See Supplemental Figures 6 and 7). Indeed, the municipalities with a higher degree of social disadvantage had a higher number of excess deaths due to COVID-19 and non-COVID-19 deaths,(28) and individuals living in municipalities with higher poverty levels had a higher risk of dying from COVID-19.(30)

Similarly, causes of death amenable to healthcare contributed to life expectancy losses between 2019 and 2021. Hospitals overwhelmed by COVID-19 were an important determinant of higher mortality in 2020(30), which very likely continued during 2021. This might be linked to the important healthcare reform that started in 2018, with the replacement of *Seguro Popular* (Popular Health Insurance) and the start of the *Instituto de Salud para el Bienestar*.(31) By 2016, 93% of the Mexican population had medical insurance,(11) while in 2022, the number of people with no access to medical insurance was estimated to be 40%.(31) Analysis of out-of-hospital deaths in Mexico during the pandemic revealed that most of the excess deaths that occurred during 2020 took place out of hospital.(32) In 2020, the regions with higher non-COVID-19 excess mortality were associated with a low percentage of the population with access to social security and healthcare.(30) Our results present a similar trend: we observed that southern Mexico reported a higher contribution of causes of death amenable to healthcare to changes in life expectancy between 2019 and 2021, while in northern Mexico, causes of death amenable to healthcare helped to mitigate life expectancy losses.

### Limitations

Our study is subject to limitations. The classification of causes of death plays an important role in our results. Some COVID-19 deaths may have been misclassified as other causes (23), particularly deaths due to acute respiratory diseases, which may have led to under-registration of deaths where COVID-19 was the true underlying cause.(28) In Mexico, the available information did not allow us to disentangle this, but we overcame this limitation by restricting our analysis to six groups of causes of death rather than individual causes. Similarly, homicides are likely to be underreported, especially in Mexico’s current militarized context. Enforced disappearances,(33) the prevalence of common and clandestine graves,(34), and the ongoing forensic crisis reflect pathways through which homicide may not be registered in vital statistics. Additionally, homicides may be improperly assigned ICD-10 codes, making them impossible to classify as such.(9)

When performing analysis at the state level in Latin American countries, we are aware of the sensitivity to the quality of the data. We used data published by the National Population Council, which has been shown to be reliable in the international context.(35) For the cause-of-death analysis, we used six broader categories rather than individual causes to minimize misclassification errors.

### Conclusion

Demographic analyses can help identify and understand structural challenges regarding population health in Mexico. Between 2015 and 2021, violence and COVID-19 were the major public health challenges that the Mexican population had to face, but the impacts of these crises have not been evenly distributed. Homicides continue to negatively impact male life expectancy, and from 2015 to 2019 they also resulted in modest declines in female life expectancy. States with lower health insurance coverage in the southern region of the country experienced higher life expectancy declines due to COVID-19 and causes amenable to medical care. Further research is necessary to better understand the sex differences in life expectancy during the first two years of the COVID-19 pandemic and to examine why life expectancy stagnated for males and continued declining for females between 2020 and 2021.

## Data Availability

All data produced are available online at

https://osf.io/cxnuh/

## Acknowledgments

The authors gratefully acknowledge the valuable suggestions from Marília Nepomuceno, Vanessa di Lego, the CED colloquium participants, and the PAA 2023 SOMEDE Session.

## Data availability statement

The data and code for this project are publicly available. The mortality data is obtained from INEGI (https://www.inegi.org.mx/programas/mortalidad/) and the population exposures from CONAPO (https://datos.gob.mx/busca/dataset/proyecciones-de-la-poblacion-de-mexico-y-de-las-entidades-federativas-2020-2070), both sources are publicly available. A complete replication package is available at OSF: https://osf.io/cxnuh/

## Authors contribution

JDZB, PV and JM conceptualized the study, JDZB performed formal analysis, MG supervised and replicated formal analysis, JDZB and JM wrote first draft, JDZB, JM, PV and MG helped write the final draft. All authors agree to submit.

## Supplementary Figures and Tables

**Supplementary Table 1.**
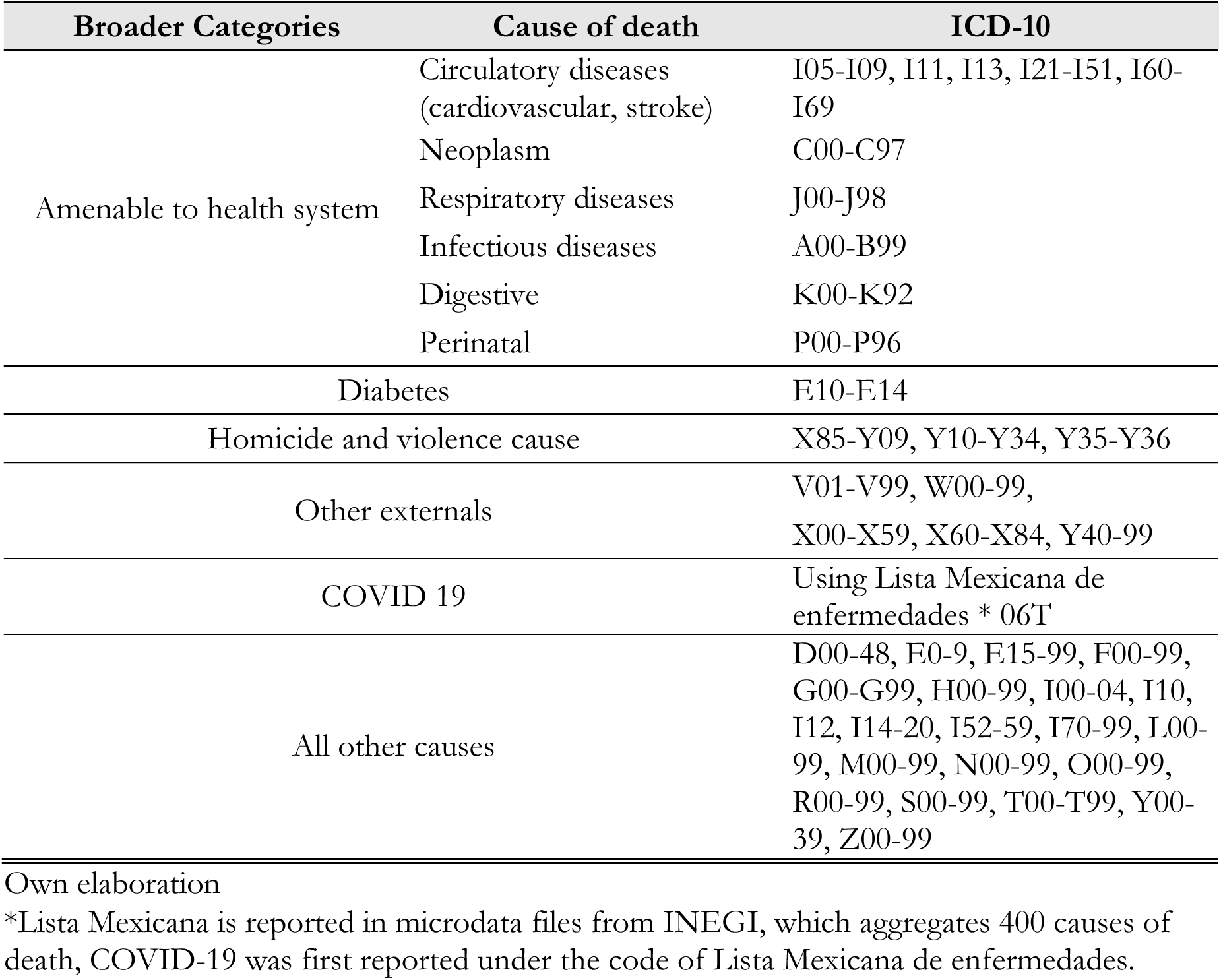
ICD codes for causes of death classification.

**Supplemental Figure 1.**
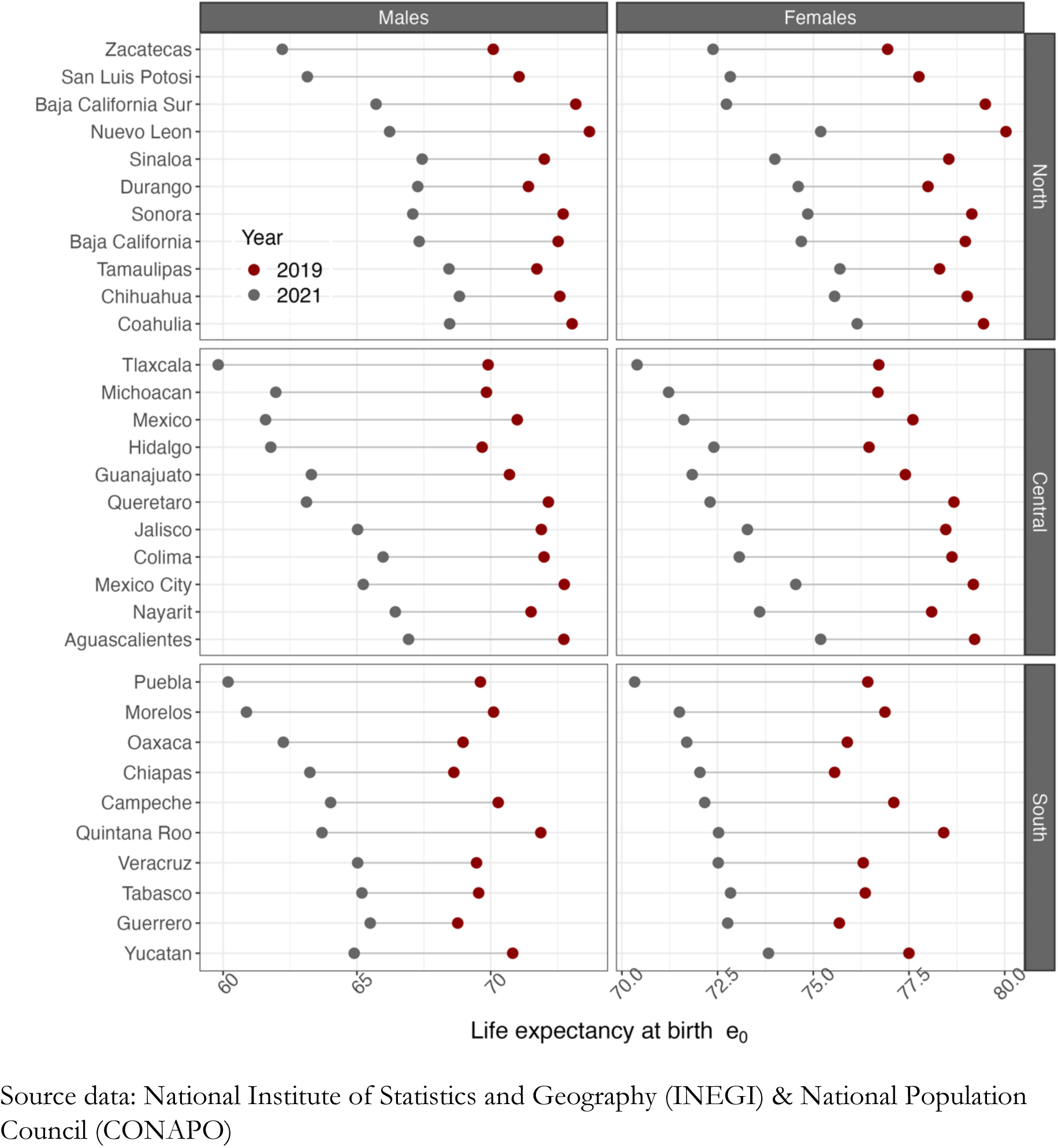
Life expectancy changes between 2019–2021, by sex, state, and region.

**Supplemental Figure 2.**
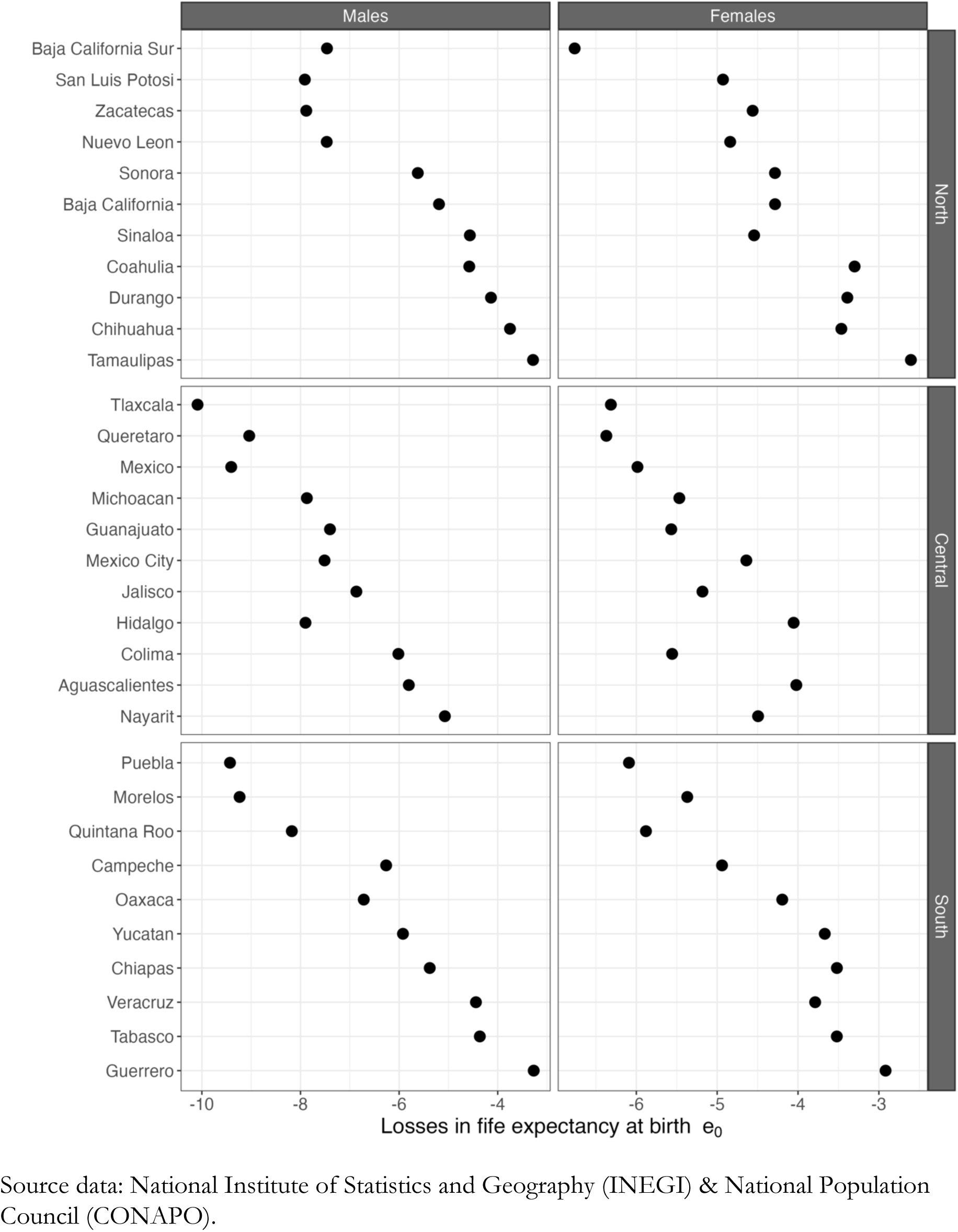
Losses in life expectancy between 2019 and 2020, across Mexican states and regions, by sex.

**Supplemental Figure 3.**
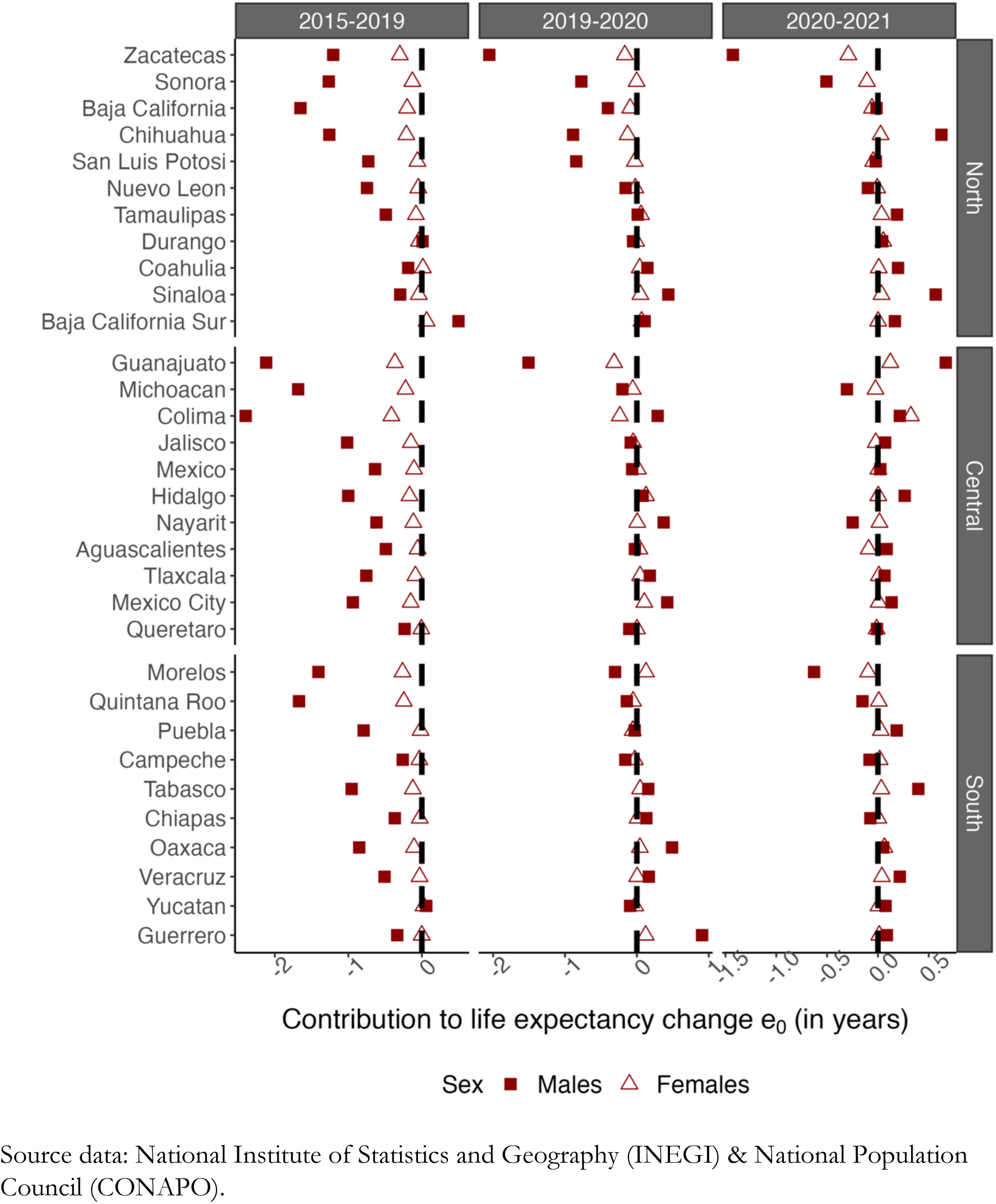
The contribution of homicides and violence to life expectancy changes in different periods by state and sex, 2015–2021.

**Supplemental Figure 4.**
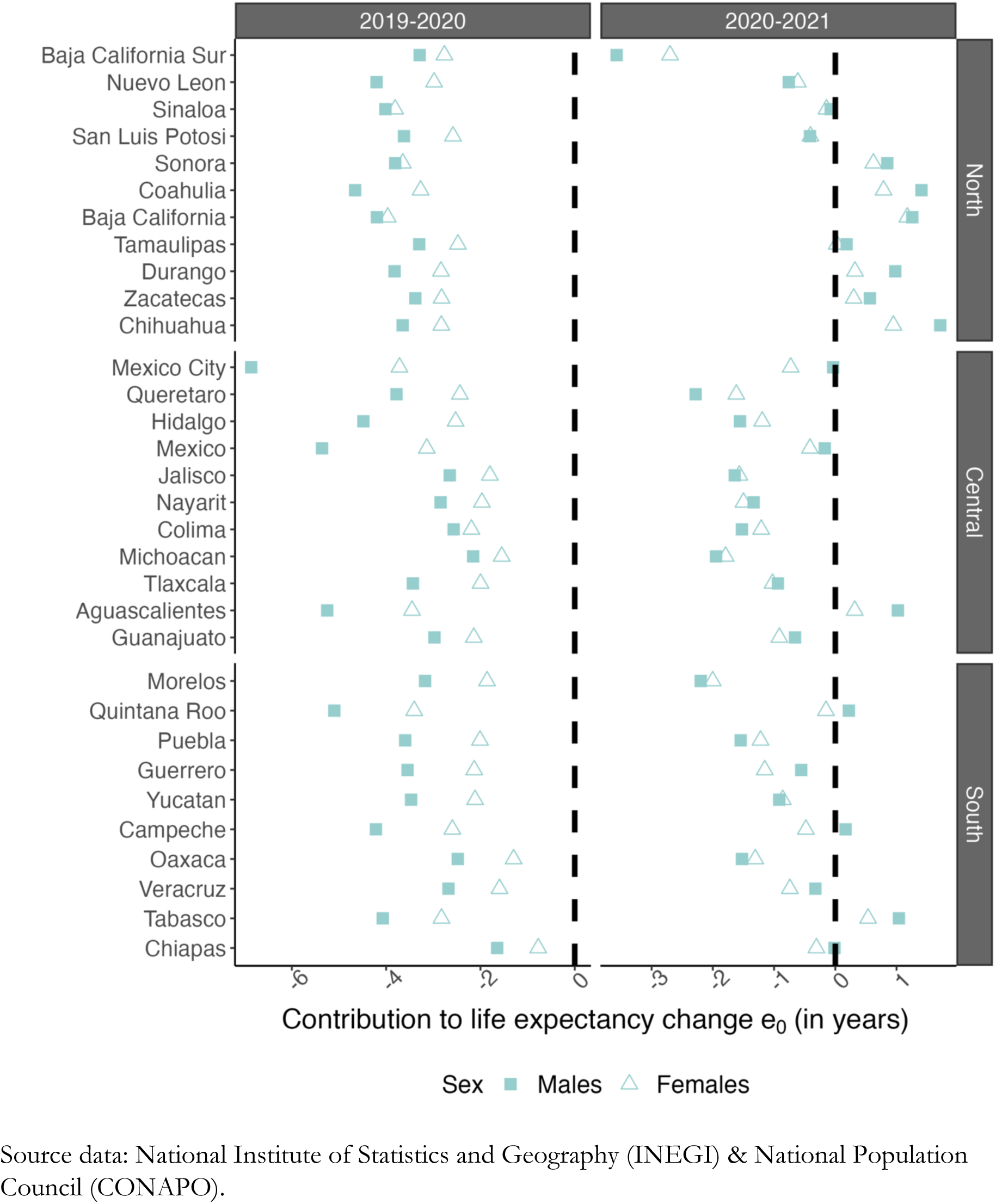
The contribution of COVID-19 to life expectancy changes in different periods by state and sex, 2019–2021.

**Supplemental Figure 5.**
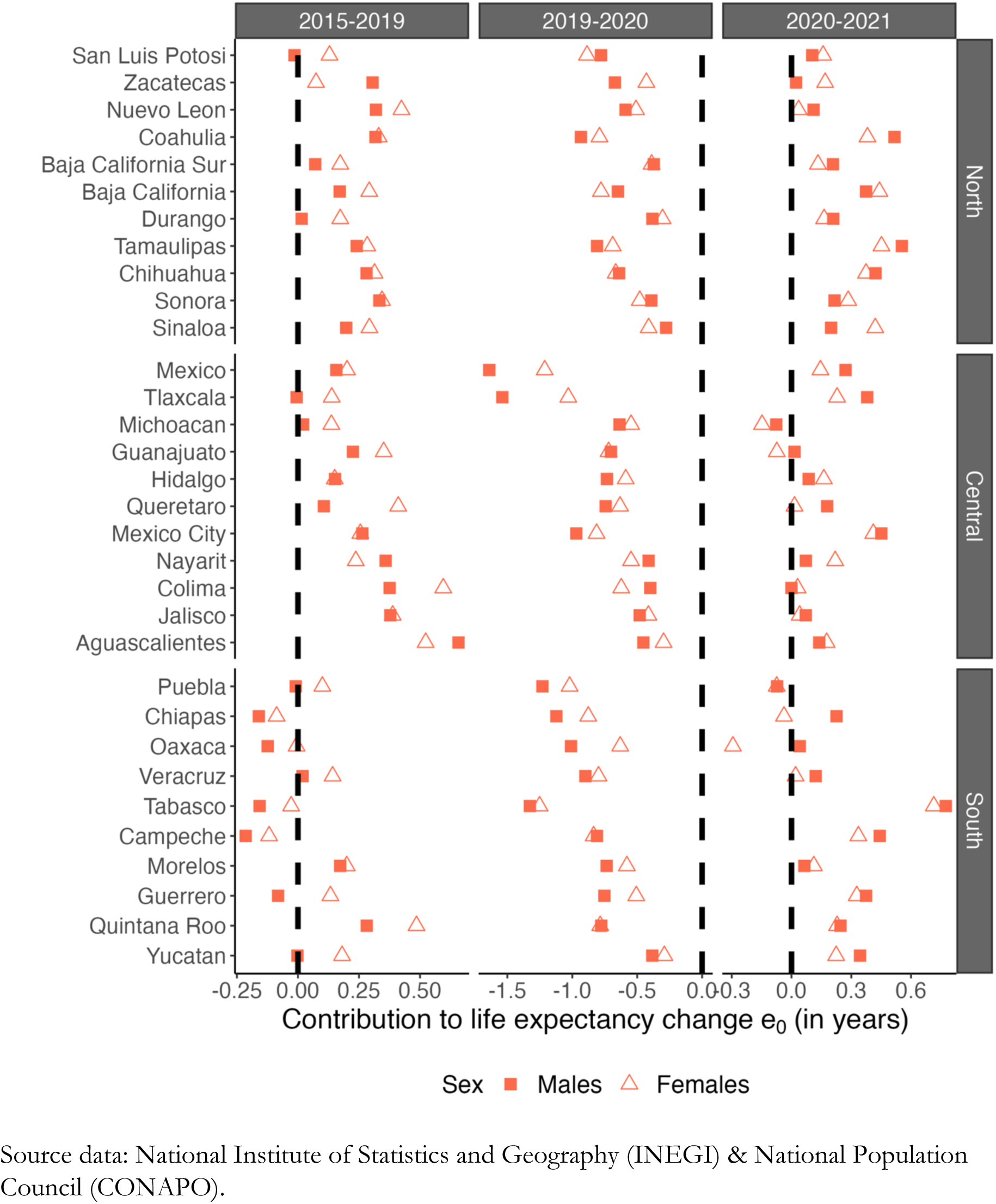
The contribution of diabetes to life expectancy changes in different periods by state and sex. Mexico, 2015–2021.

**Supplemental Figure 6.**
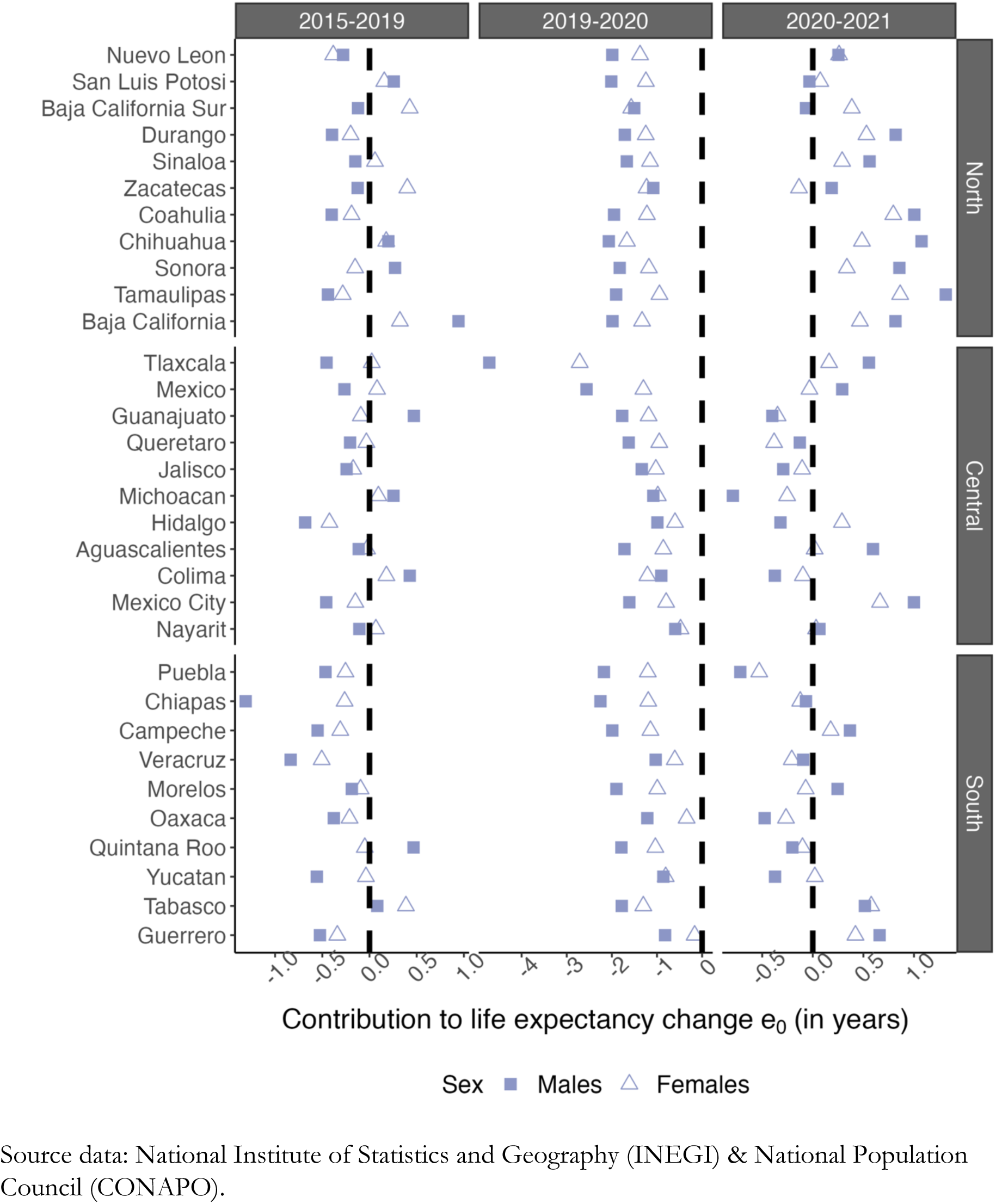
The contribution of causes amenable to healthcare to life expectancy changes in different periods by state and sex, 2015–2021.

**Supplemental Figure 7.**
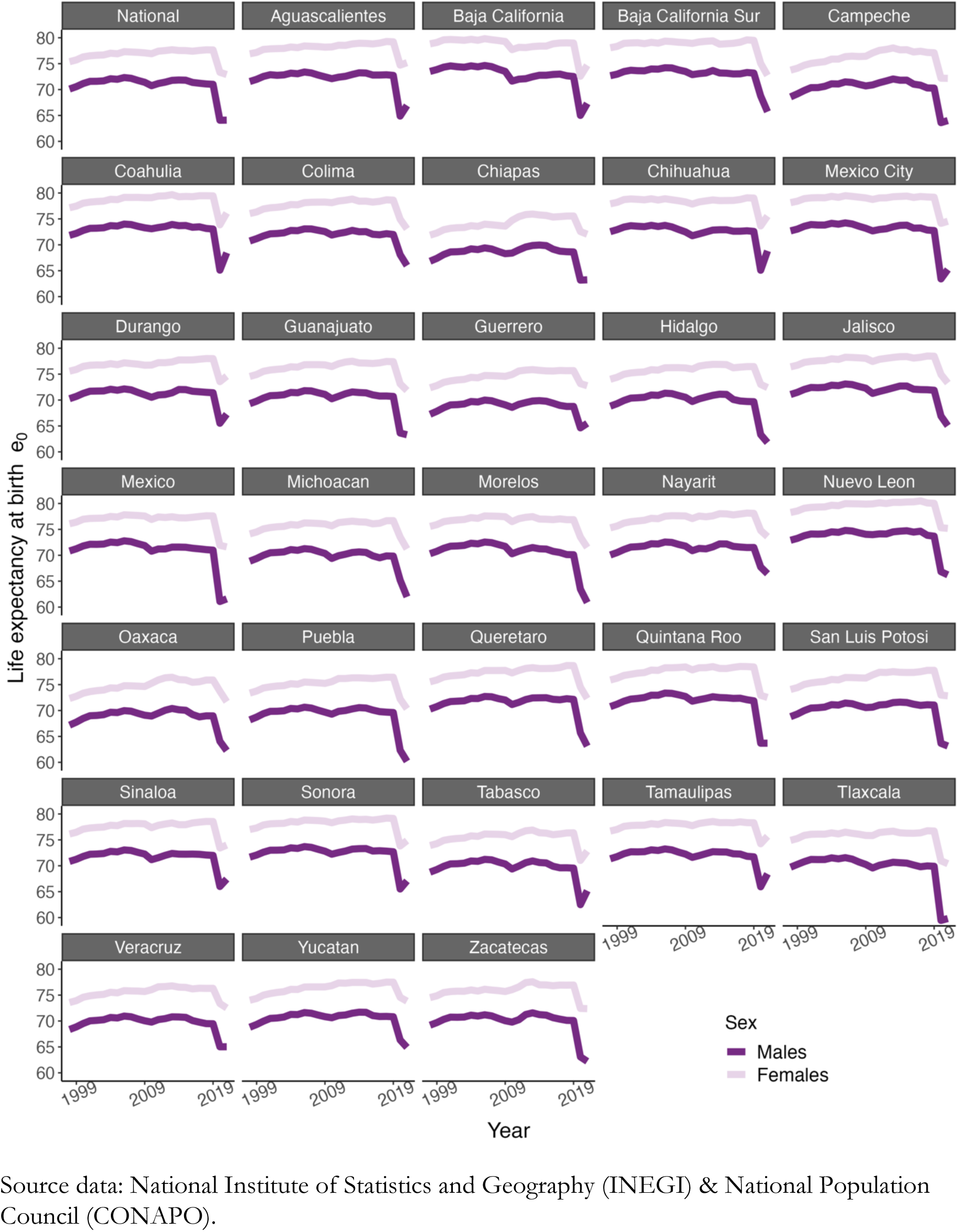
Time trends in life expectancy at birth across Mexican states and the national level, by sex. Mexico, 1999-2021.

**Supplemental Figure 8.**
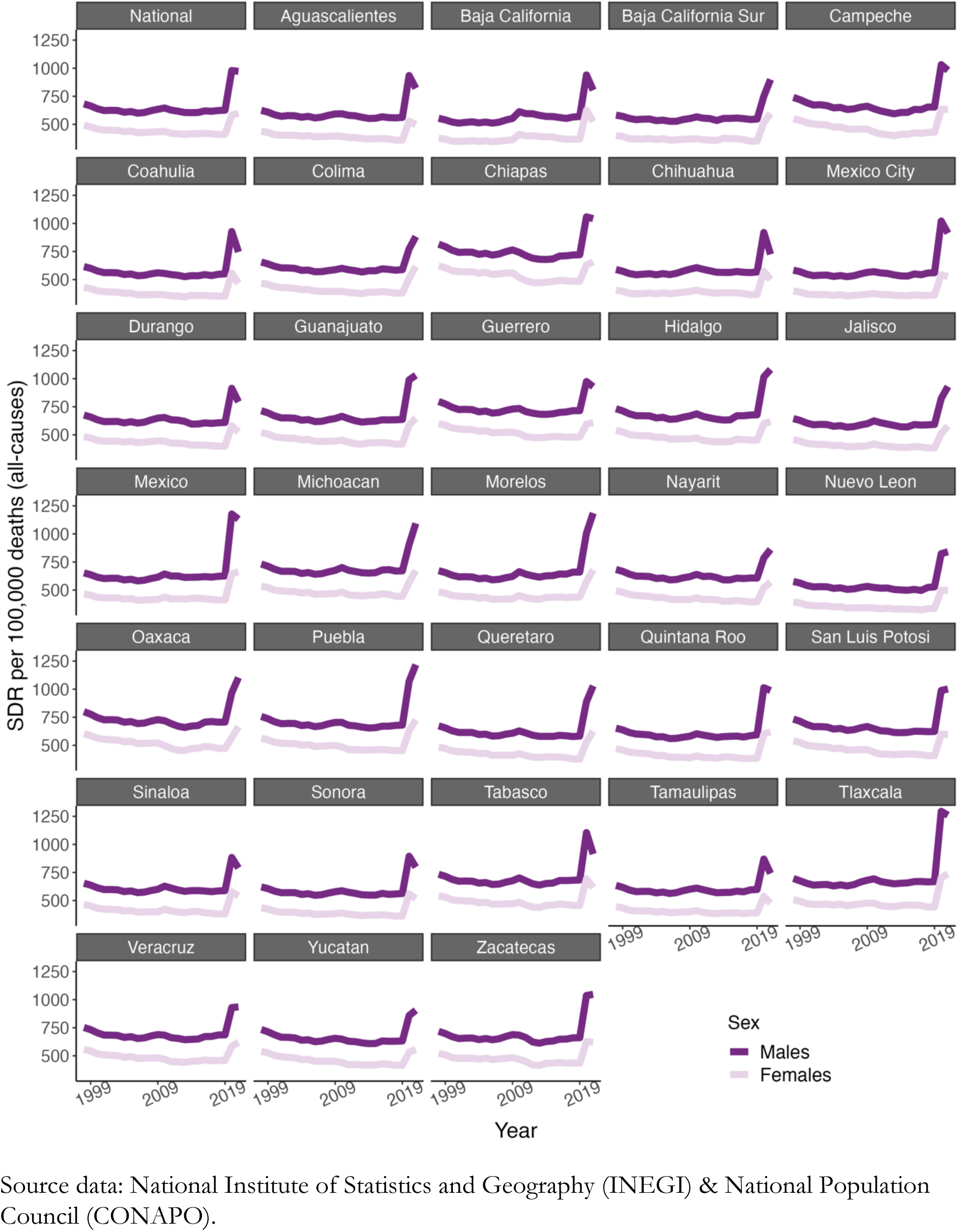
Time trends in age-standardized death rates per 100,000 inhabitants (all causes) across Mexican states and the national level, by sex. Mexico, 1999-2021.

**Supplemental Figure 9.**
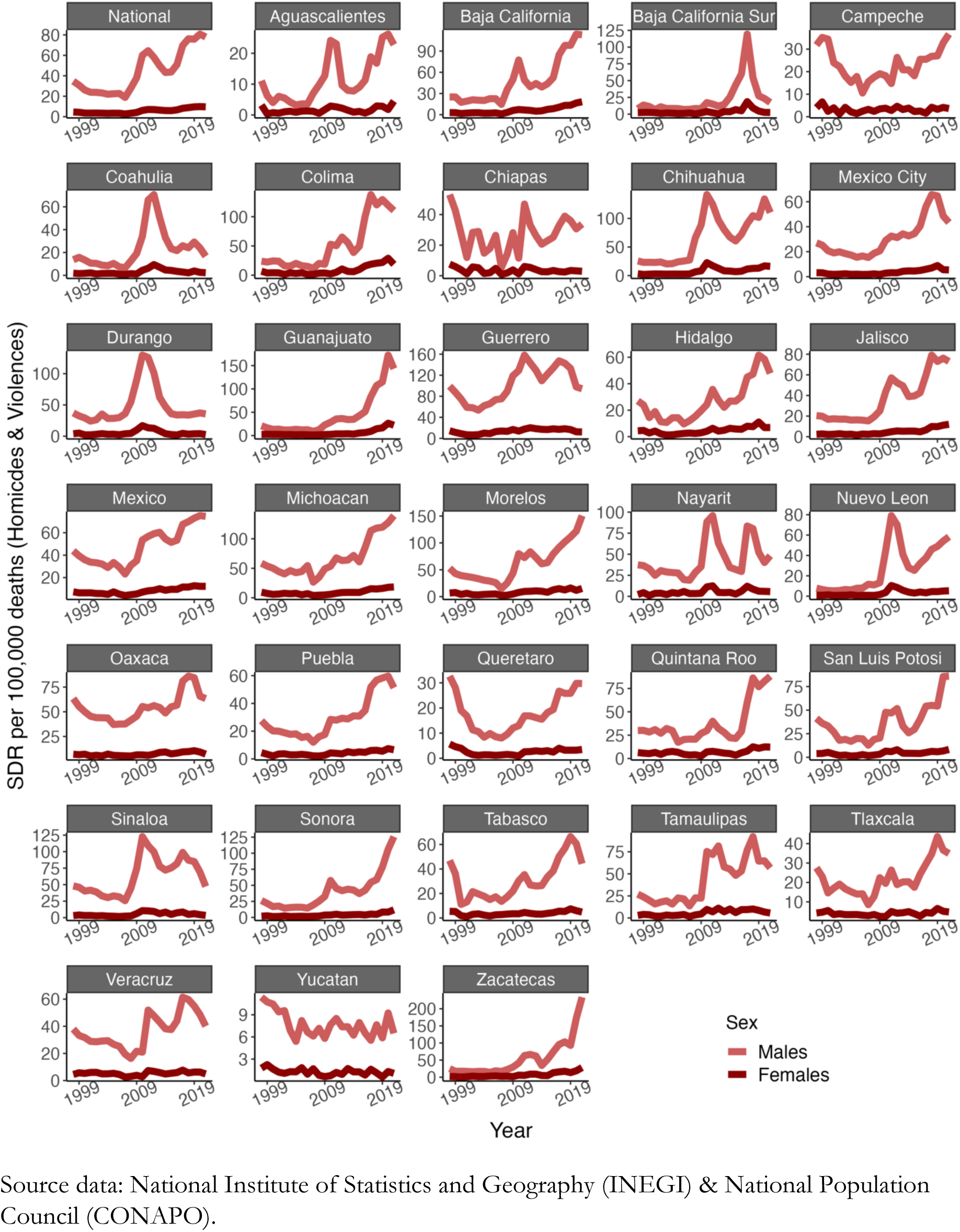
Time trends in age-standardized death rates per 100,000 inhabitants (homicides and violent deaths) across Mexican states and the national level, by sex. Mexico, 1999-2021.

**Supplemental Figure 10.**
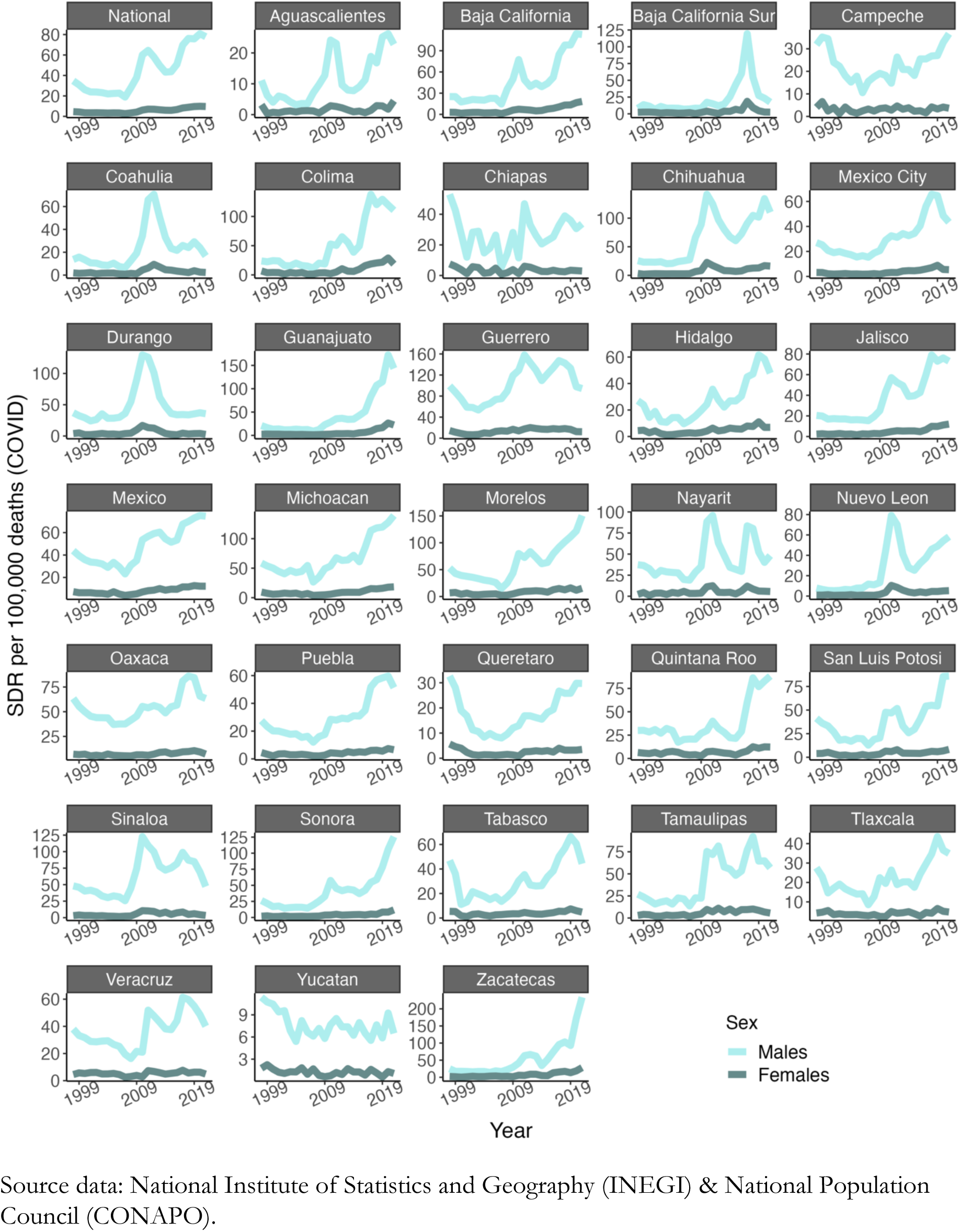
Time trends in age-standardized death rates per 100,000 inhabitants (COVID-19) across Mexican states and the national level, by sex. Mexico, 1999-2021.

**Supplemental Figure 11.**
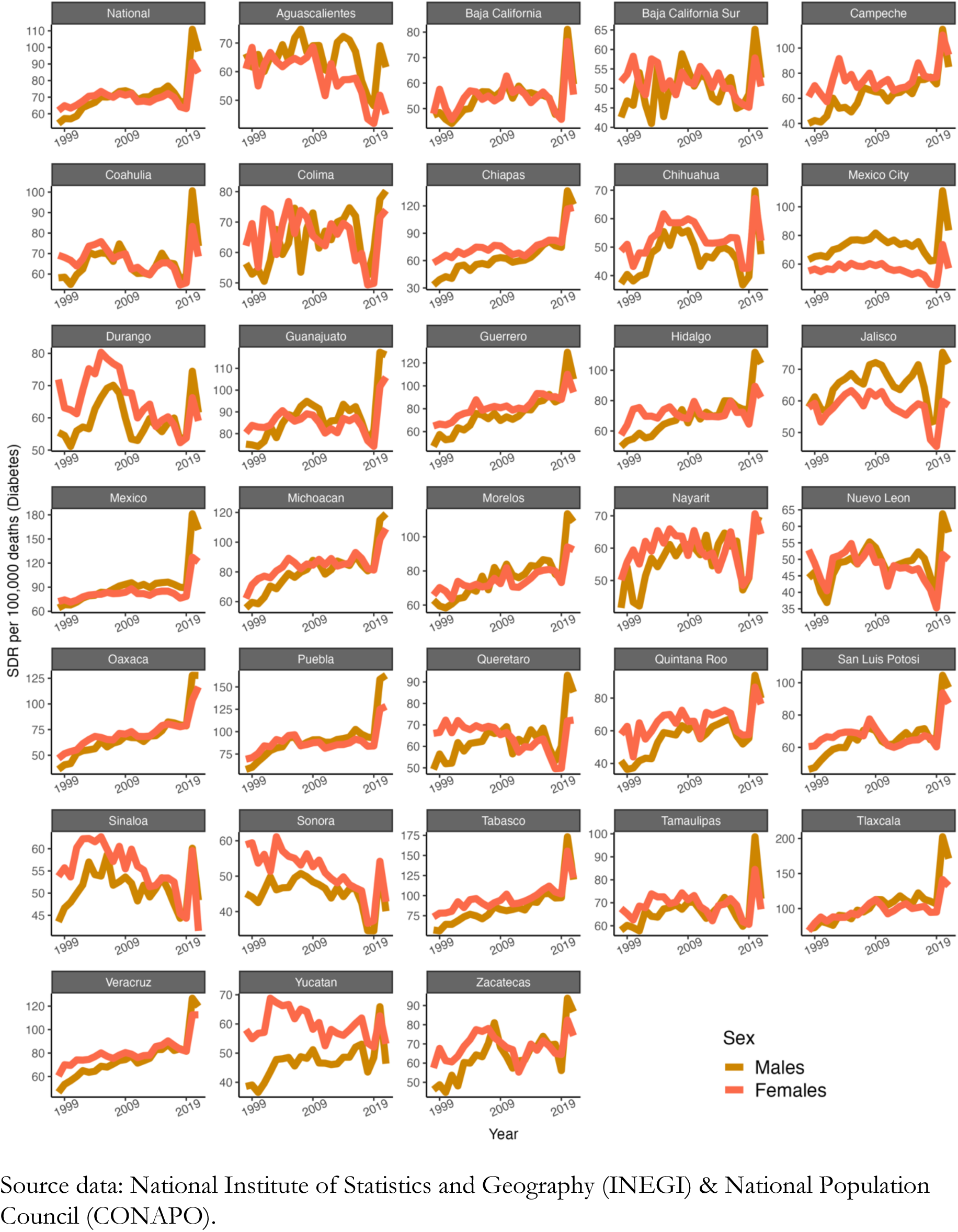
Time trends in age-standardized death rates per 100,000 inhabitants (Diabetes) across Mexican states and the national level, by sex. Mexico, 1999-2021.

**Supplemental Figure 12.**
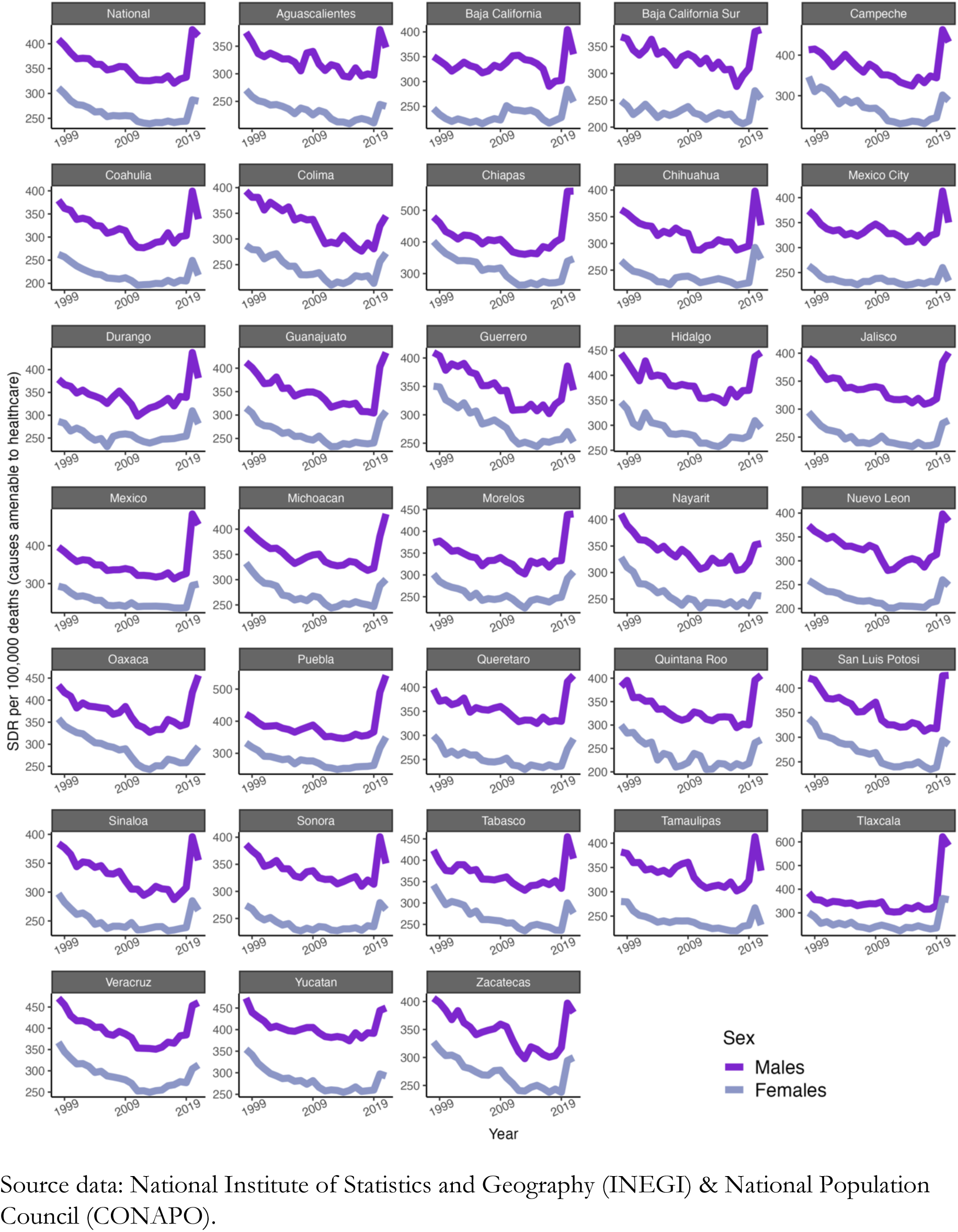
Time trends in age-standardized death rates per 100,000 inhabitants (causes amenable to healthcare) across Mexican states and the national level, by sex. Mexico, 1999-2021.

**Supplemental Figure 13.**
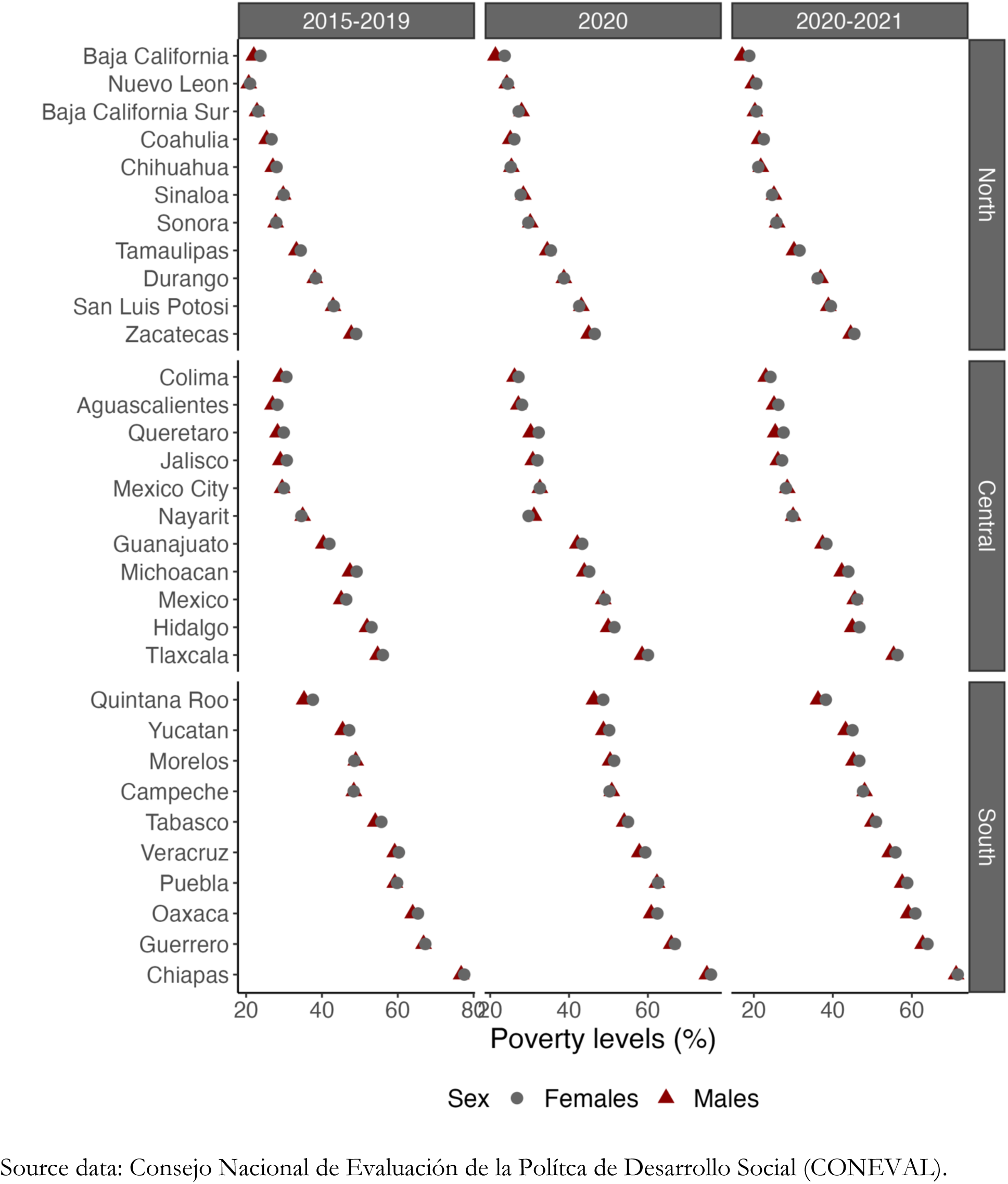
Time trends in age-standardized death rates per 100,000 inhabitants (causes amenable to healthcare) across Mexican states and the national level, by sex. Mexico, 1999-2021.

**Supplemental Figure 14.**
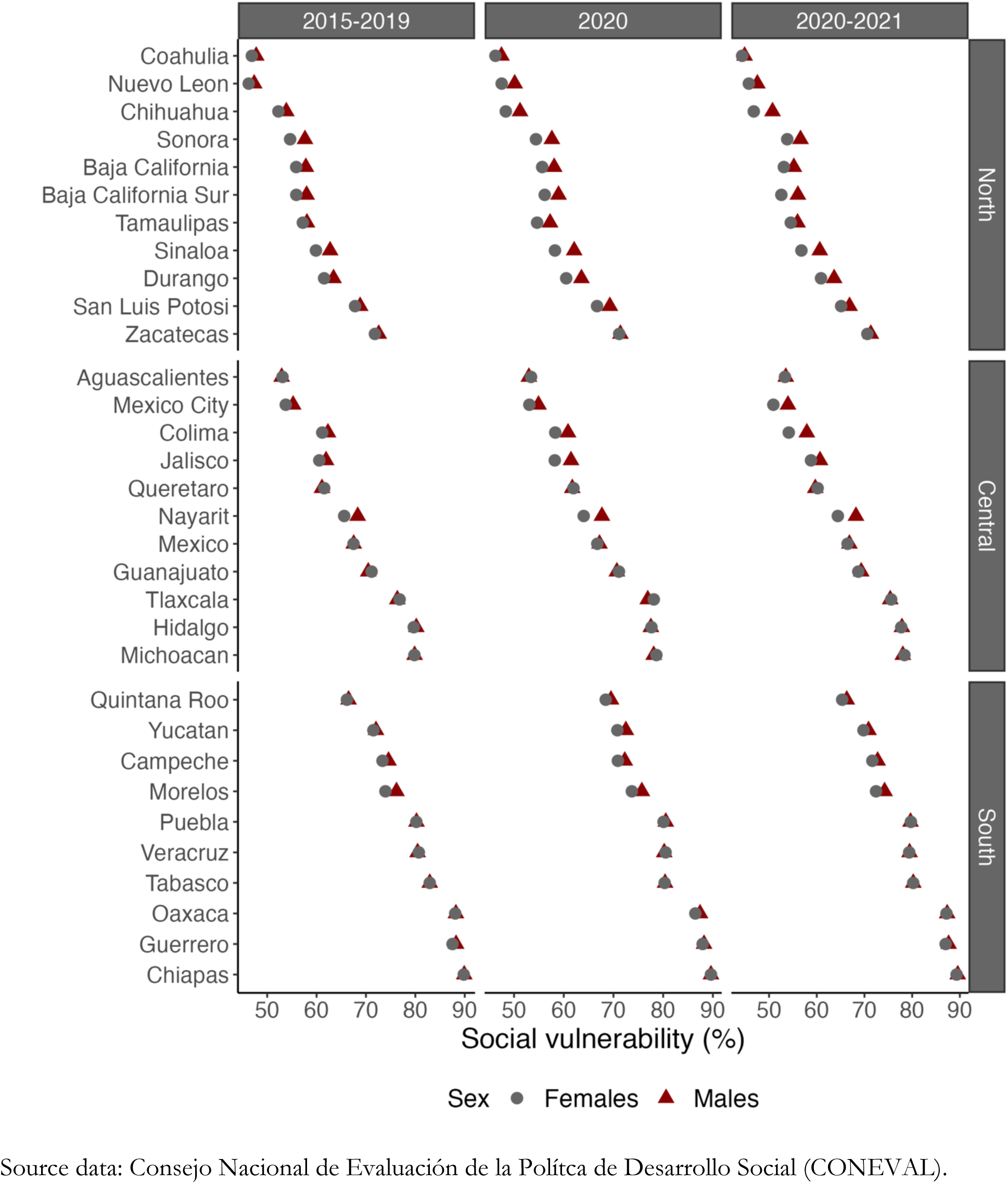
Time trends in age-standardized death rates per 100,000 inhabitants (causes amenable to healthcare) across Mexican states and the national level, by sex. Mexico, 1999-2021.

**Supplemental Figure 15.**
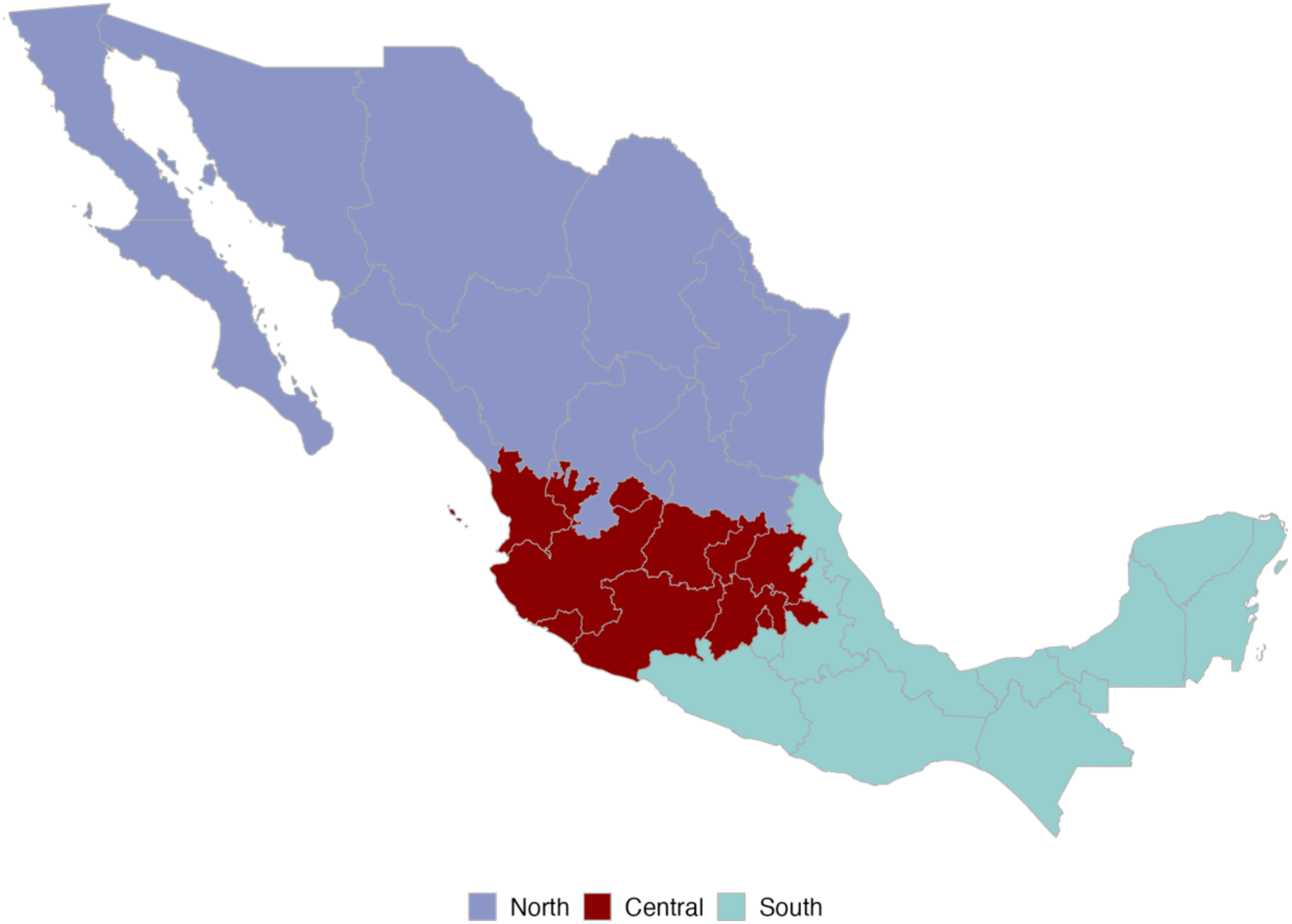
Map of regions in México.

